# Incorporating Mass Vaccination into Compartment Models for Infectious Diseases

**DOI:** 10.1101/2022.04.26.22274335

**Authors:** Glenn Ledder

## Abstract

The standard way of incorporating mass vaccination into a compartment model for an infectious disease is as a spontaneous transition process that applies to the entire susceptible class. The large degree of COVID-19 vaccine refusal, hesitancy, and ineligibility, and initial limitations of supply and distribution require reconsideration of this standard treatment. In this paper, we address these issues for models on endemic and epidemic time scales. On an endemic time scale, we partition the susceptible class into prevaccinated and unprotected subclasses and show that vaccine refusal/hesitancy/ineligibility has a significant impact on endemic behavior, particularly for diseases where immunity is short-lived. On an epidemic time scale, we develop a supply-limited Holling type 3 vaccination model and show that it is an excellent fit to vaccination data. We also extend the Holling model to a COVID-19 scenario in which the population is divided into two risk classes, with the highrisk class being prioritized for vaccination. For both cases with and without stratification by risk, we see significant differences in epidemiological outcomes between the Holling vaccination model and naive models. Finally, we use the new model to explore implications for public health policies in future pandemics.

## 1. Introduction

As we have seen with COVID-19, vaccination is a critical component of the public health response to pandemics, particularly those caused by novel diseases. Models can help us integrate vaccination and mitigation practices into a coherent public health policy by showing what happens when mitigation practices are relaxed early in the vaccine rollout phase. They can also help us assess the relative benefits of proposed vaccination programs. However, meaningful modeling results depend on a realistic accounting for vaccination in an epidemiological model. A lot of work has been done on vaccination programs (see [22], for example), but the incorporation of vaccination into models remains problematic. The purpose of this paper is to contribute to the development of better vaccination models, beginning with models for mass vaccination.

Mass vaccination is generally incorporated into dynamical system disease models as a single-phase spontaneous transition process that ultimately moves everyone out of the susceptible class who does not contract the disease first [2, 4, 25]. These assumptions are questionable on two grounds: first, the combination of ineligibility because of age or physical condition, hesitancy that delays vaccination or causes people to receive only a partial course, and outright refusal, all of which have impacted vaccination against COVID-19, suggests that vaccine non-acceptance* needs to be incorporated into infectious disease models; second, the dynamics of the single-phase spontaneous transition does not match the dynamics of vaccination during the rollout of a new vaccine.

In a single-phase spontaneous transition, the rate is proportional to the size of the leaving class. This assumption is mathematically convenient, but it is often biologically inaccurate. In a true spontaneous transition, such as radioactive decay, the absolute rate of transition in a collection of individuals is largest at the beginning, when the collection itself is largest. This is not true for disease processes, such as recovery, where a mean time of 5 days means that most transitions occur in days 4, 5, and 6. The modeling error caused by using a spontaneous transition for recovery in an endemic disease model is small because the overall rate at steady state is dependent primarily on the mean time and not the distribution of times. For an epidemic model, we often assume spontaneous transitions in spite of the modeling error; alternatively, we could use a multi-phase transition model, corresponding to an Erlang distribution of times [1, 19]. In this way, we can control the standard deviation of the transition time as well as the mean. For more control over the shape of the distribution, we could use an integral equation formulation [13].

For vaccination in a model for a novel disease, the problem with assuming spontaneous transition is different in character. Those of us who wanted to be vaccinated as soon as a vaccine was approved found that we could not get our shots until our risk class was cleared; even then, the waiting time for an appointment was sometimes long. A spontaneous transition model imposes no limit to the number of transitions that can occur simultaneously, whereas in vaccination there is a hard limit to the number of appointment slots, one which is initially low because of supply limitations and saturates to the capacity of the administrative system. A better model will need to account for both limited supply and limited distribution capacity.

The issues raised here play out differently depending on the time scale of the model. Problems of supply and distribution are important on an epidemic time scale of weeks to months, but not on an endemic time scale of years. For endemic models, the primary concern with the standard treatment of vaccination is vaccine non-acceptance, which probably will not abate significantly over time. We examine this issue in Section 2 by considering the impact of vaccine non-acceptance in a variant of the standard SIR endemic model that incorporates vaccination, vaccine non-acceptance, and loss of immunity.

On an epidemic time scale, it is less clear that vaccine non-acceptance needs to be considered. As long as people who want to be vaccinated are waiting for their turn, the existence of people who will not be vaccinated should not matter. On the other hand, both supply and distribution problems should matter. We deal with these issues in Section 3 by introducing several models that incorporate supply and distribution limitations. These models are parameterized using vaccination data from the United States and then tested using a variant of the standard SEIR epidemic model that incorporates vaccination and vaccine non-acceptance.

The vaccination model of Section 3 is extended in Section 4 to a COVID-19 scenario, in which individuals are divided into multiple risk classes, with vaccine allocation based on a protocol that favors those believed to be at high risk for hospitalization. We conclude this section with an exploration of the impacts of improvements in vaccine acceptance, vaccine manufacture speed, and mitigation practices.

## 2. Vaccine Non-Acceptance in an Endemic Disease Model

Figure 1 shows a schematic that incorporates vaccination, vaccine non-acceptance, loss of immunity, and demographic processes of birth and death into a relatively simple disease model. The class structure is PUIRU, a modification of the SIRS model [15] in which the susceptible class is subdivided into (P)revaccinated and (U)nprotected subclasses. We assume a constant total population *N* = 1, achieved via a uniform death rate of *µY* for each class *Y*, with no disease-induced deaths, and compensated by a total birth rate *µ*. Prevaccinated and unprotected individuals become infected at rates *βPI* and *βUI*, respectively, and prevaccinated individuals move into the removed class by vaccination at rate *ϕP*. Infectious individuals are removed at rate *γI*. The influx of susceptibles from birth is partitioned into fractions *a* + *r* = 1, where *a* is the fraction of individuals who accept vaccination. The fate of removed individuals depends on whether they are vaccine acceptors or non-acceptors. We assume that vaccine acceptors will renew their immunity when booster doses are recommended; hence, they stay in the removed class. Non-acceptors lose immunity at rate *θR*. The special cases of full vaccine acceptance and no loss of immunity are achieved by setting *r* = 0 and *θ* = 0, respectively.

**Figure 1.**
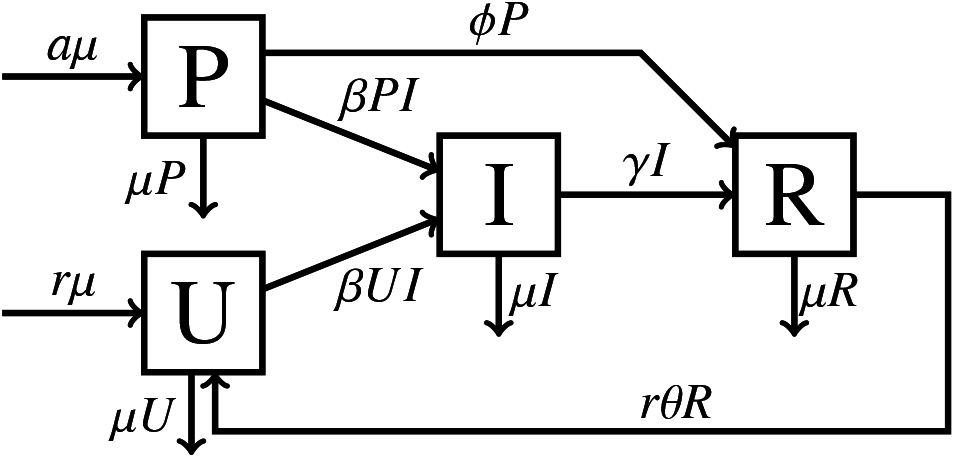
The PUIRU endemic model.

The model assumes that vaccine-induced immunity occurs in 100% of vaccinations and lasts as long as disease-induced immunity. Neither assumption is true for COVID-19 [12], but they are reasonable simplifying assumptions, given the broad goal of the model. The results for this model will serve as an upper bound on the effectiveness of vaccination for a real disease.

While the standard way of writing the model would be to use the mutually exclusive classes P, U, I, and R, it is more convenient^†^ to think of the model as having mutually exclusive classes S, I, and R, where *S* = *P* + *U* is the total susceptible population fraction, along with an additional state variable P.

Thus, we have differential equations

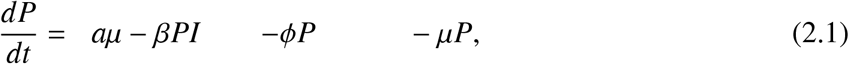

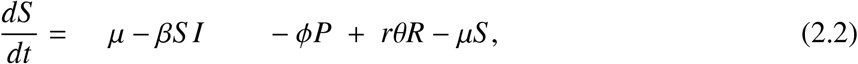

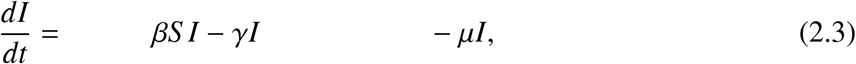

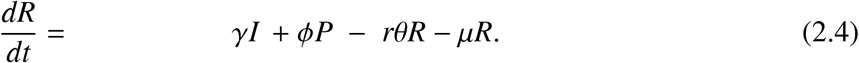

To simplify the analysis, we scale the system using the mean lifespan 1*/µ* as the time scale,^‡^ resulting in the model (with equations reordered to facilitate use of the next generation method for the basic reproduction number)

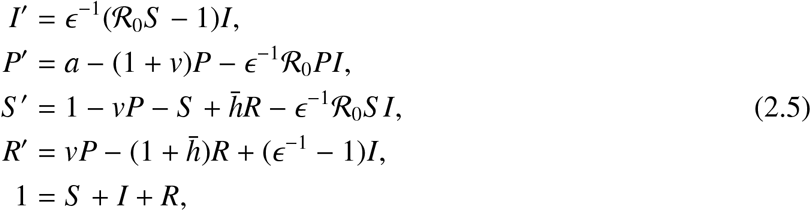

where the prime symbol refers to the derivative with respect to scaled time and the new dimensionless parameters are

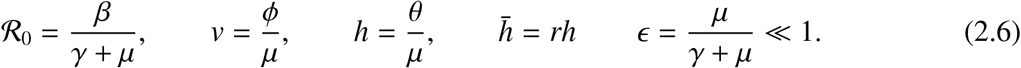

Note that ℛ_0_ is the basic reproduction number in the absence of vaccination. The parameter *v* represents the ratio of expected lifespan to the expected time required to vaccinate a class P individual. The parameter *h* represents the expected number of times a non-accepting individual loses immunity during their lifespan, while 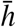 is the expected number of immunity losses in a lifespan irrespective of acceptance. Finally, *ϵ* is the ratio of the mean time in class I to the mean lifespan. Since human infections last weeks while a normal lifespan is measured in years, the numerical value of *ϵ* is small enough that terms of *O*(*ϵ*) can be neglected when added to terms of *𝒪*(1).

To prepare for this asymptotic analysis, we rescale the model using

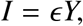

thus obtaining the final version:

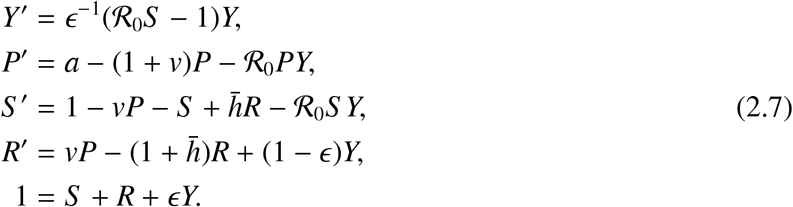

### 2.1. The Disease-Free Equilibrium and the Vaccine-Reduced Reproduction Number

The disease-free equilibrium is characterized by *Y* = 0, which decouples the equilibrium equations. Of principle interest is the disease-free total susceptible population fraction, which is

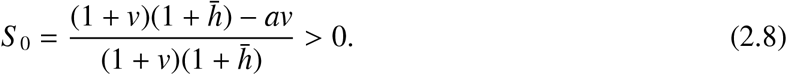

This quantity is needed to compute the reproduction number in the presence of vaccination (which we will call the ‘vaccine-reduced reproduction number’),

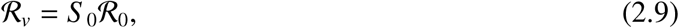

which can be found using the next generation method [2, 30].

### 2.2. Stability Analysis of the PUIRU Model

As with most simple endemic disease models, the equilibrium and stability properties of the model are simple, albeit tricky to prove. We summarize the result in a theorem and defer the proof to an appendix.

**Theorem 1**. *The PUIRU model (2.7) has the following properties:*

1. *There is a unique disease-free equilibrium, with susceptible population fraction given by (2.8)*. *This equilibrium is asymptotically stable if and only if* ℛ_*v*_ < 1.
2. *There exists an endemic disease equilibrium if and only if* ℛ_*v*_ *>* 1. *This equilibrium is unique, with infectious population fraction*

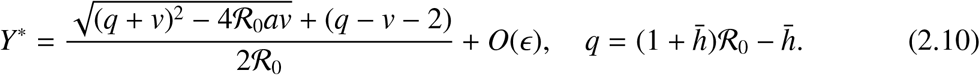
3. *The endemic disease equilibrium is asymptotically stable*.

### 2.3. Effect of Vaccine Non-Acceptance on Herd Immunity

Achievement of herd immunity is a principal outcome that can be used to measure the success of a public health vaccination program (see [26], for example). For the PUIRU model, herd immunity requires ℛ_*v*_ < 1, or

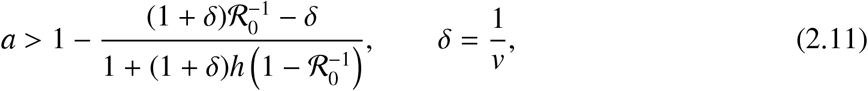

where we have introduced *δ* = 1*/v* because we expect *v* to be large. Taking *δ* = 0 provides an upper bound for the effectiveness of the vaccine.

The results are displayed in Figure 2 as contours showing the combination of disease characteristics that corresponds to herd immunity for a given acceptance level, with *v* → ∞. With an optimistic acceptance level of *a* = 0.9, herd immunity can be reached with ℛ _0_ < 10, provided there is no loss of immunity. As immunity loss increases, the results change quickly, with requirements ℛ _0_ < 4 with *h* = 2, ℛ _0_ < 3 with *h* = 3.5, and ℛ _0_ < 2 with *h* = 8. For a highly-infectious disease with short-lived immunity, like COVID-19, herd immunity is not a practical goal.

**Figure 2.**
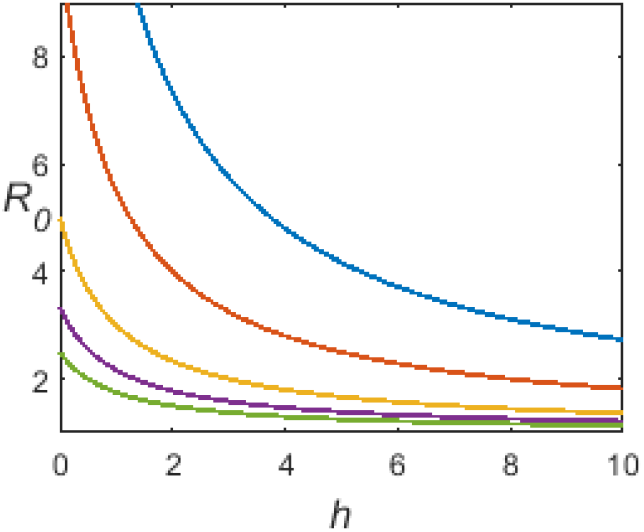
Disease characteristics for herd immunity for given acceptance: *a* = 0.95, 0.9, 0.8, 0.7, 0.6, upper right to lower left, with *v* → ∞.

### 2.4. Effect of Vaccine Non-Acceptance on Infection Prevalence

If the vaccine-reduced reproduction number is large enough for the endemic disease equilibrium to be stable, then the most important measure of vaccine impact is the relative decrease in the average infectious population [27], which is

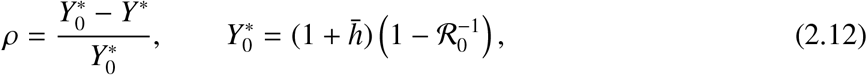

where 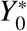 is the rescaled infectious population fraction when *v* = 0. In the limit *v* → ∞, which serves as an upper bound of vaccine impact, this is

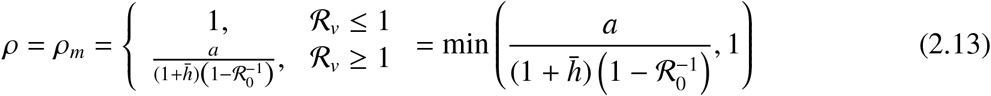

The results for maximum vaccine impact *ρ*_*m*_ are shown in Figure 3, using ℛ_0_ values of 2, 4, and 8 for the left, center, and right panels. Within each panel are curves for *h* = 0, 1, 3, 9, 30 from top to bottom. When the conditions for herd immunity are not met, vaccine non-acceptance causes a significant reduction in vaccine impact. The loss due to non-acceptance is limited for disease with no loss of immunity, but increasing rate of immunity loss carries with it a much greater penalty for vaccine non-acceptance. For diseases with a very high rate of immunity loss and a modest or large basic reproduction number (the bottom curves of the center and right panels), such as COVID-19, a non-acceptance fraction of 30% reduces the impact of the vaccine by 90% or more. This is particularly problematic because the vaccine fatigue caused by the need for frequent boosters can be expected to increase non-acceptance.

**Figure 3.**
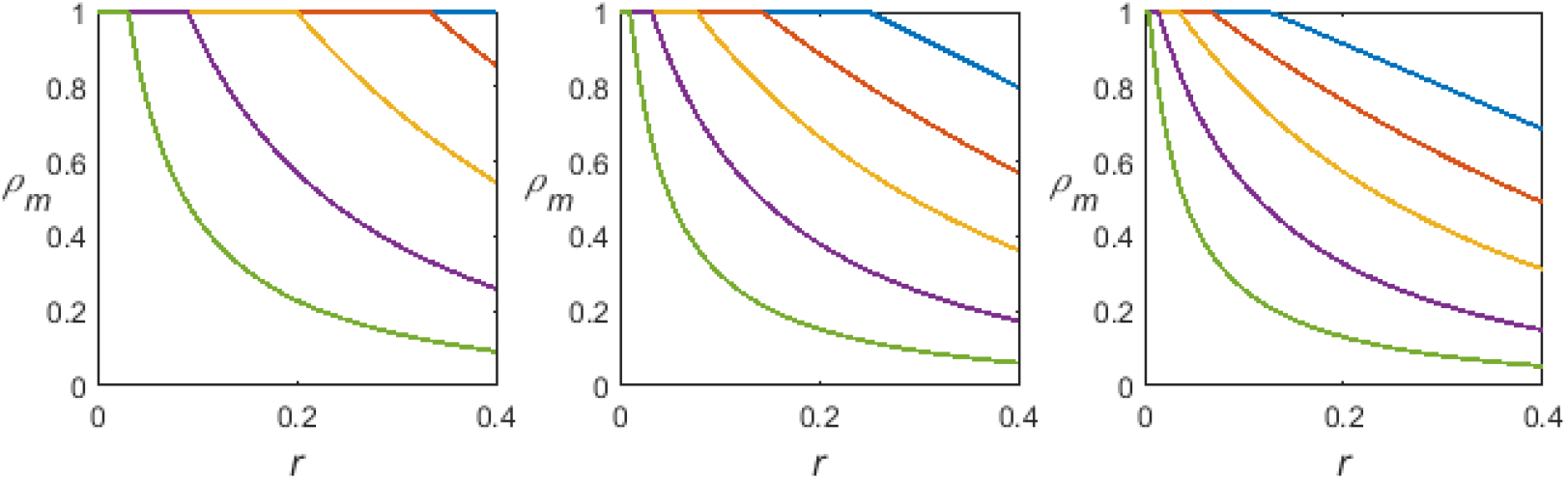
The ratio *ρ*_*m*_ that marks an upper bound for vaccine impact; ℛ_0_ = 2, 4, 8 for left, center, and right panels, respectively, and *h* = 0, 1, 3, 9, 30 from top to bottom in each panel.

## 3. Vaccination Models with Limited Distribution and Supply

As noted in the introduction, the standard single-phase spontaneous transition model for vaccination fails to account for limited supply and distribution. Here we develop some models that address this flaw. These models do not account for loss of immunity and administration of booster doses. By the time these factors are important, we should not need to worry about the issues of limited supply and distribution capacity; hence, a model similar to the endemic model of Section 2 would be appropriate.

One complicating factor in modeling vaccination over short time scales is the administration as a two-dose regimen rather than as a single dose in those countries whose vaccination programs are based on mRNA vaccines. In the context of an epidemiology model, the two-dose regimen adds additional classes, which is a complication unlikely to be important enough to affect overall results. Based on the crude assumption that one dose of mRNA vaccine gives half the protection of the two-dose regimen, we simply count each dose administered as one-half of a vaccination.

### 3.1. Model Development

As before, let *a* be the fraction of people who will accept vaccination. With *V*(*t*) as the fraction of the population that has been vaccinated (half the number of doses administered since the beginning of vaccination relative to the total population) and *W*(*t*) as the fraction of the population that is still waiting for vaccination, we have

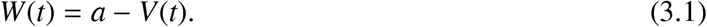

Our vaccination model will focus on the epidemiological class *W*, but we can use the model with (3.1) to compute *V* for parameter fitting against data.

We assume that the coefficient in the spontaneous transition rate is a saturating function of time as the vaccine supply increases; thus,

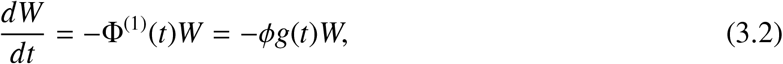

where Φ^(1)^ is the relative vaccination rate and 0 < *g*(*t*) ≤1 is a nondecreasing function that accounts for limited supply of vaccine doses during the initial phase of the rollout. The simplest assumption is that supply increases linearly up to its maximum; that is,

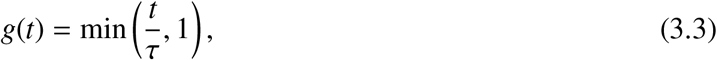

where *τ* is the number of days required to achieve the maximum.

There is good reason to think that this first model will be inadequate. While it accounts for limited supply, it fails to account for limited distribution capacity. For the special case where supply is not limited, we should expect the initial vaccination rate to be largely independent of the number of people waiting to be vaccinated, as it would be limited instead by the number of available vaccination appointments per day. In terms of the process, it is reasonable to think of vaccination as analogous to enzyme kinetics and predation. There is a large pool of patients (substrate/prey) and a finite number of vaccinators (enzyme molecules/predators). Initially, each vaccinator spends nearly all of its time processing patients. Only when the size of the patient pool becomes relatively small do the vaccinators have to spend time searching for patients. This description leads to the supply-adjusted MichaelisMenten/Holling type 2 model,

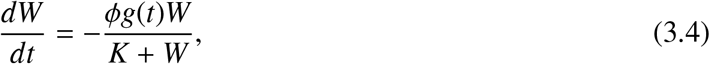

with *ϕ* the theoretical maximum rate of vaccination (‘achieved’ as *W* →∞ and *g* = 1, except that in actuality *W* ≤ 1), and *K* the value of *W* for which the rate is half of the theoretical maximum. In the context of an epidemic model, it will be convenient to partition this formula into factors as

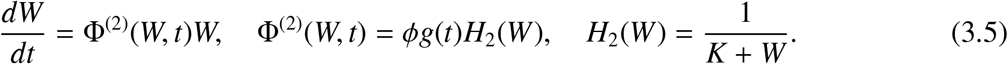

We should also consider the Holling type 3 model,

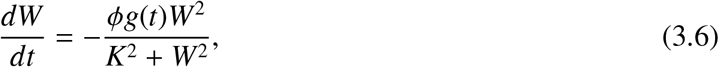

or

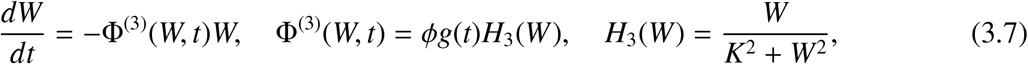

which accounts for the observation in some predator-prey systems that searching can be less efficient when the target population is very low.

Figure 4 shows the fits of the three models to data for the United States [6, 29], with the standard spontaneous transition with full vaccine acceptance as a control. The time period of December 20, 2020 through July 7, 2021 runs from the date the vaccine was authorized for general use to the date at which booster doses were authorized. The best-fit parameters (using two significant figures unless the first digit is 1) are

**Figure 4.**
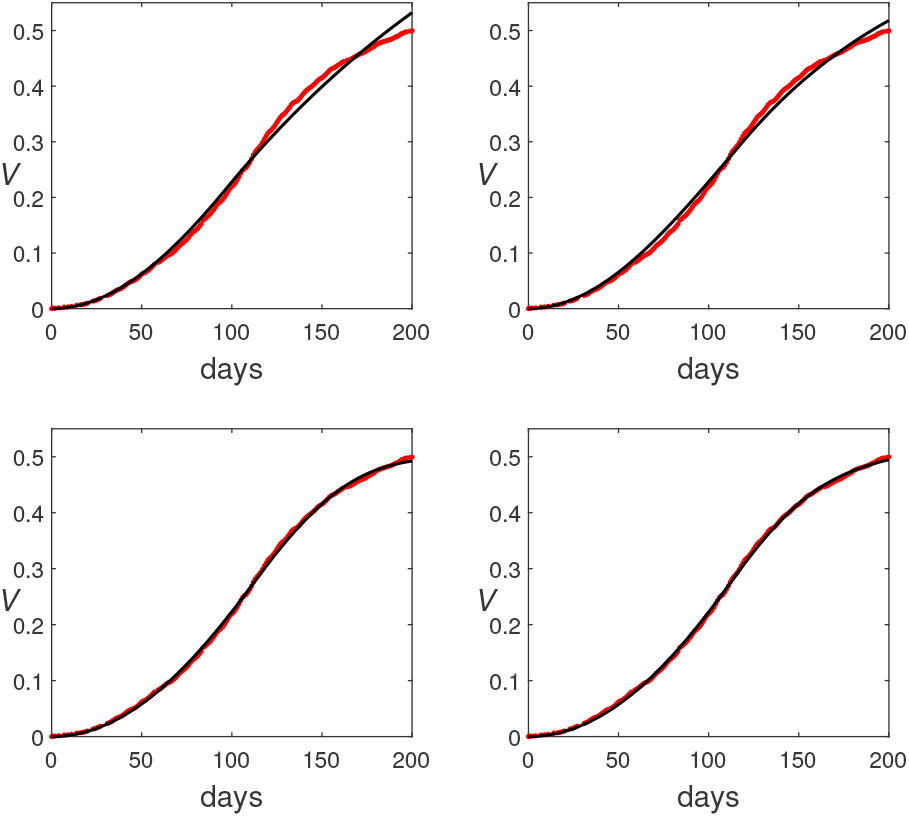
Four supply-limited vaccination models (black) with data for vaccination in the United States from December 20, 2020 through July 7, 2021 (red): top left, (3.2) without vaccine non-acceptance; top right, (3.2); bottom left, (3.4); bottom right, (3.6).

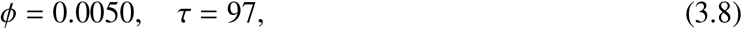

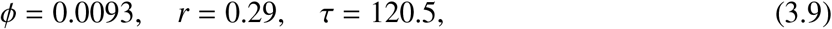

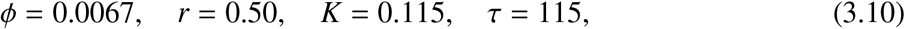

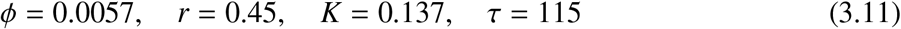

for the linear (3.2) with full vaccine acceptance, linear with vaccine non-acceptance, type 2 (3.4), and type 3 (3.6) models, respectively. The linear model with full vaccine acceptance is a poor match for the data. The linear model with non-acceptance is better, but it clearly shows a systematic error relative to the data and the non-acceptance fraction *r* = 0.29 is objectively too low in comparison to the data. The Holling models show an excellent fit by visual standards. They are almost identical except for a slight difference near the end of the period, when the type 2 model levels off faster than the type 3. The AIC values for the four models are -1634, -1707, -2069, and -2146, respectively, a difference large enough to identify the type 3 model as the best on empirical grounds. Of course the parameters would need to be determined separately for each country, and it is possible that the type 2 model might be better for some. However, the type 3 assumption of slow search rate when the target population is low seems appropriate, so we are justified in preferring type 3 on mechanistic grounds as well as empirical.

### 3.2. Vaccination in an SEIR Model

To incorporate vaccination into the standard SEIR epidemic model,^§^ we split the susceptible class into prevaccinated and unprotected subclasses, as in Section 2 (see Figure 5). For the simplest epidemic model, we neglect processes that occur on a demographic time scale—birth, death, and loss of immunity. We also assume that prevaccinated and unprotected individuals have the same encounter rates, the same probability of encounters with infected individuals, and the same probability of being infected by an encounter with an infected individual. For simulations, the initial susceptible population is divided into the P and U subpopulations, each of which subsequently decreases through infection, while the P subpopulation also decreases through vaccination.

**Figure 5.**
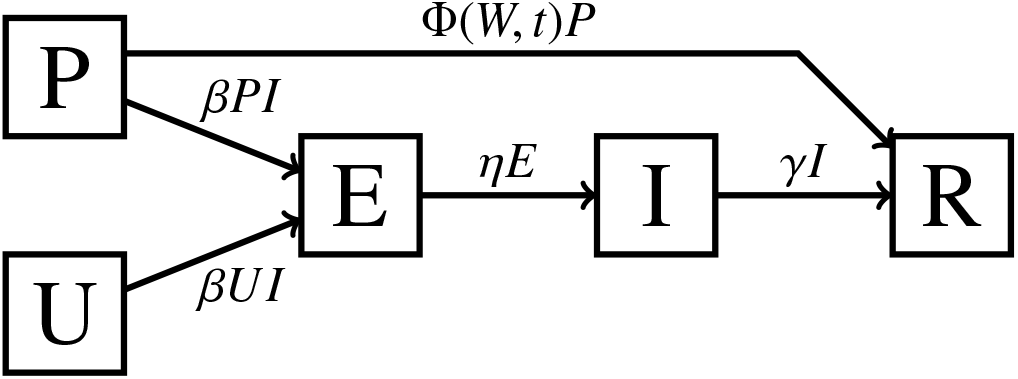
The PUEIR epidemic model.

We assume (as was the case with COVID-19 in the United States and most other countries) that vaccination is administered without regard for possible prior immunity; hence, a fraction *P/W* of vaccinations are administered to individuals in the prevaccinated class, which is why the function Φ is the same for the epidemiological model as for the corresponding vaccination model. The full model simply couples the differential equation for *W* with the epidemic model of the figure,

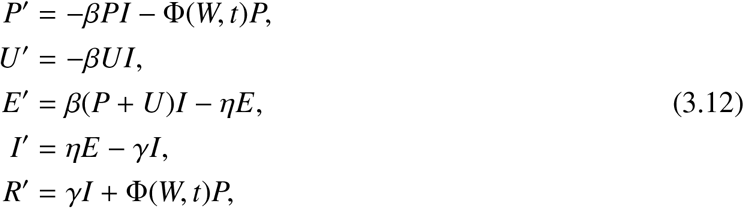

using any desired vaccination rate function Φ.

Figure 6 shows the result using the Holling type 3 vaccination model (3.7) with best-fit parameters in a scenario with initial conditions of 30% immunity, 2% infectious, and 1% exposed, with an effective reproduction number (incorporating any mitigation policies along with the unmitigated basic reproduction number) of 3. The key outcome is that vaccination comes too slowly to make much of a difference. The rate of new infections hits its maximum at around day 35, with barely 5% of the population already vaccinated. The effective end of the outbreak occurs around day 75, at which point vaccination has not reached even 20% of the population. The main benefit of vaccination is to decrease the susceptible class for the next outbreak.

**Figure 6.**
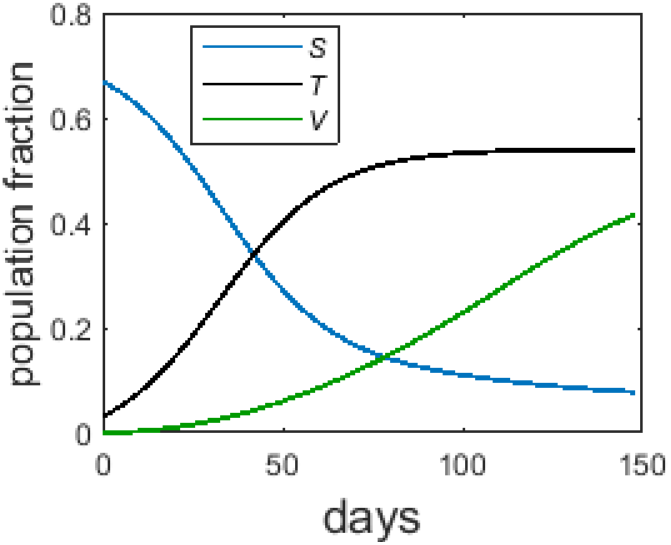
The PUEIR model with Holling type 3 vaccination (3.7, 3.12). Parameters are *γ* = 0.1, *η* = 0.2, ℛ_0_= 3, and vaccination parameters from (3.11), with initial conditions *R*(0) = 0.3, *I*(0) = 0.02, *E*(0) = 0.01, *P*(0) = 0.67*a, U*(0) = 0.67*r. S* = *P* + *U* is the total susceptible fraction and *T* is the cumulative fraction infected, given by *T* ^′^ = *β*(*P* + *U*)*I, T* (0) = *E*(0) + *I*(0).

To see what difference the choice of vaccination model makes, we consider two scenarios, shown in Figure 7. We take the same disease parameters of *γ* = 0.1, *η* = 0.2, and ℛ_0_ = 3, along with the best fit values for *r* and *K* for each model. The upper panels assume best fit values for the maximum vaccination rate constant *ϕ* and the time to maximum *τ*, while the lower panels assume that changes in vaccine research and public health funding allow for a doubling of the maximum rate constant and a reduction of the time to maximum from the 2021 values to 30 days. Initial conditions are *R*(0) = 0.3,^¶^ *I*(0) = 0.1, *E*(0) = 0.05, with the remaining population split into *P* and *U* according to the best fit non-acceptance parameter *r*.

**Figure 7.**
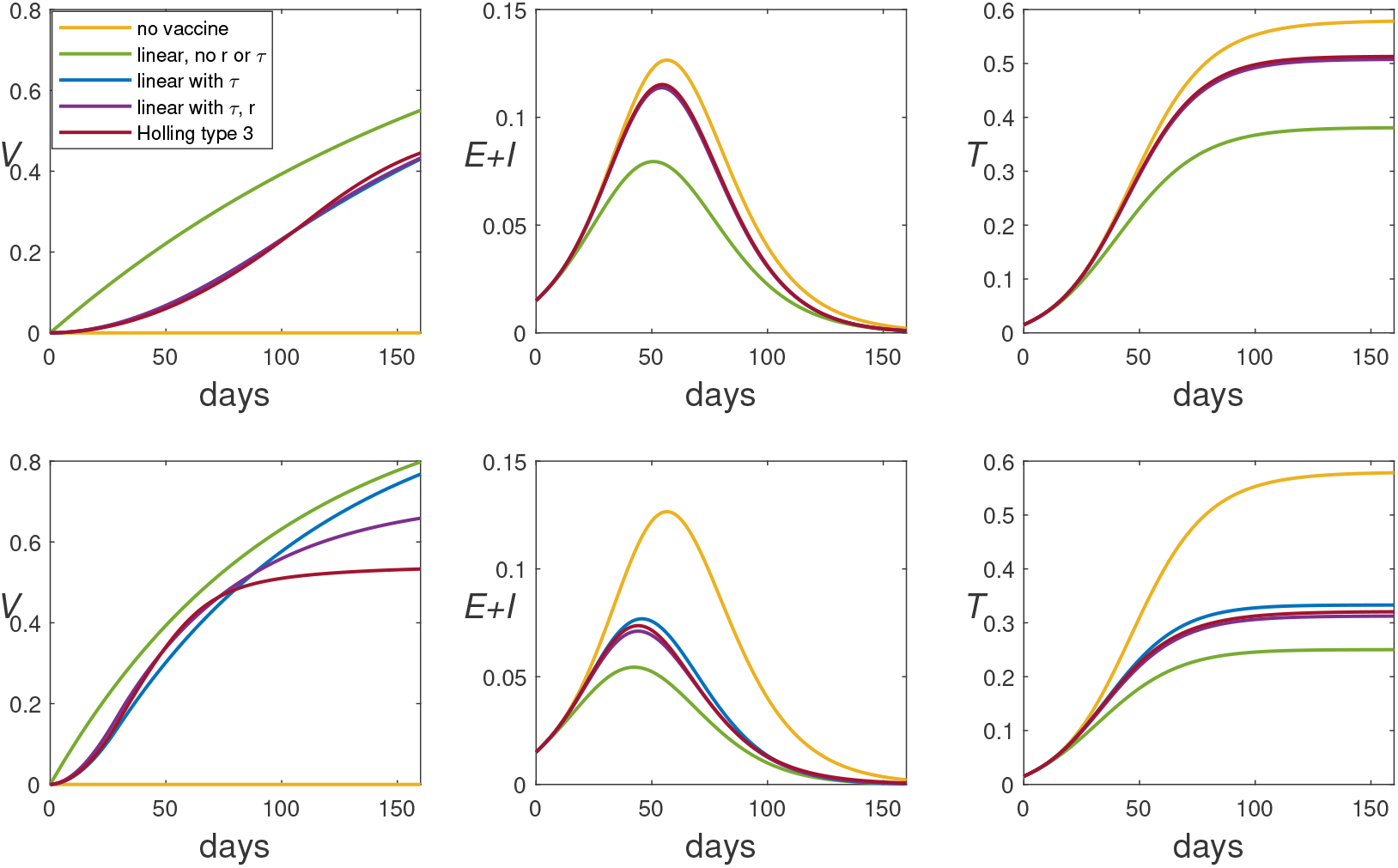
Comparison of vaccination models for two scenarios, both with *γ* = 0.1, *η* = 0.2, ℛ_0_ = 3, *R*(0) = 0.3, *I*(0) = 0.1, *E*(0) = 0.05, and best fit values for *r* and *K*; the top panels assume best fit values for *ϕ* and *τ*, while the latter assume twice the best-fit values for *ϕ* and 30 days for *τ*. The left panels show the fraction of the population that has been vaccinated, the middle panels show the current fraction that is infected, and the right panels show the cumulative fraction infected.

The top panels show only slight differences between the three models that account for limited supply; that is, vaccine non-acceptance (seen in the two lower curves in the legend) and limited distribution capacity (seen in the bottom curve in the legend) make only a slight difference in the epidemiological outcomes. In the more optimistic scenario of the lower panels, the noticeable difference in the vaccination outcome has some effect on the epidemiological outcomes. In both scenarios, the incorporation of any vaccination into the models clearly affects the model outcomes, while naive vaccination models that fail to account for supply limitations, as represented by the model labeled ‘linear, no *r* or *τ*,’ give epidemiological results that are far too optimistic. Independent of the choice among the three supply-limited vaccination models, the greater supply in the lower scenario clearly makes a large difference in the epidemiological outcomes.

## 4. A COVID-19 Vaccine Rollout Scenario

Suppose we want to model the introduction of vaccine for COVID-19. The PUEIR model of the previous section is too simple to capture important details of COVID-19. Specifically, we have to be able to account for three different levels of disease—asymptomatic, mild symptomatic, and prehospitalized symptomatic—and we have to be able to account for public health measures such as social distancing, testing, and isolation.

Incorporating vaccination into an earlier SEAIHRD COVID-19 model [20] is more complicated than merely splitting the S class into P and U subclasses. Because COVID-19 involves different levels of risk depending on age and health status, the administration of the vaccine prioritized individuals according to predefined classes.^*^ This feature of vaccination needs to be incorporated into a COVID19 epidemiological model.

### 4.1. A Model for COVID-19 with Two Risk Classes

The compartment diagram of Figure 8 represents a model for the scenario that occurred when vaccination first became available for COVID-19. The classes are Prevaccinated, Unprotected, Exposed (latent), Asymptomatic, (symptomatic) Infectious, Hospitalized, Removed, and Deceased. Subscripts 1 and 2 are used for low and high risk subclasses for P, U, and E and for standard symptomatic vs prehospitalized subclasses for I. The model incorporates the following assumptions:

1. Susceptible individuals, either prevaccinated or unprotected, become infected at a relative rate proportional to an ‘effective infectivity’ count *X*. Where the infectious class I is the only class capable of transmitting the disease in an SEIR model, the COVID-19 model has different categories of infectives with different levels of infectivity, which contribute to the effective infectivity in different ways (see below). Public health measures require additional detail.
2. Low-risk latent individuals (*E*_1_) become infectious with relative rate constant *η*; a fraction *p* of these become asymptomatic, while the remainder become symptomatic. Similarly, high-risk latent individuals become infectious with relative rate constant *η*, but here a fraction *q* become prehospitalized and the remainder symptomatic.^†^
3. Asymptomatic individuals and mildly symptomatic infectious individuals (*A* and *I*_1_) gradually recover with relative rate constants *α* and *γ*, respectively. Prehospitalized infectives become hospitalized with relative rate constant *σ*.
4. Hospitalized individuals leave the hospital with relative rate constant *ν*; a fraction *m* of these die while the rest recover.
5. Vaccination moves low-risk prevaccinated individuals into the removed class, with relative rate constant that depends on the status of the full vaccination (*W*) and high-risk vaccination (*W*_2_) programs, as described below. Vaccination moves high-risk individuals into the low-risk unprotected class (*U*_1_) with probability *s*, or the removed class.^‡^
6. Recovered individuals are immune for long enough that we can ignore possible loss of immunity.
7. Deaths from unrelated causes and births are sufficiently small over the course of the scenario that they can be ignored.
8. Mixing of individuals is homogeneous; that is, high-risk and low-risk individuals have the same total encounter rate and the same probability of encounters being with high-risk individuals.

**Figure 8.**
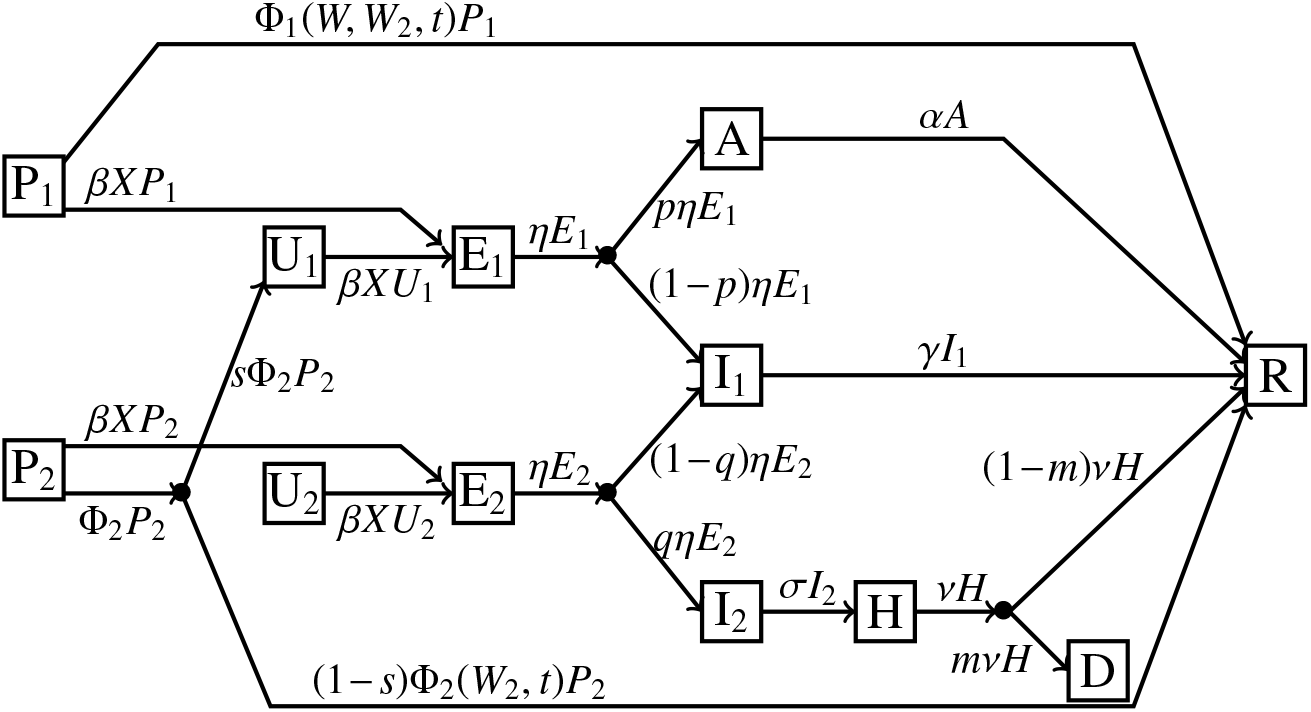
The PUEAIHRD epidemic model.

Several of the assumptions are oversimplifications, but ones that are regularly made and justifiable in epidemiology models whose purpose is to obtain general results rather than to fit data. For example, incorporating loss of immunity would add extra detail by requiring the model to distinguish between vaccinated individuals and individuals with disease-induced immunity; however, little would change because most vaccination occurs late in the 200-day scenario. Also, a more realistic heterogeneous model for population mixing would likely have some impact on the results [10], but would require additional assumptions regarding the nature of the mixing. Specific results might change with more detail in the model, but the broad patterns of how vaccination and the way it is modeled impact results should be the same.

The model consists of the differential equations

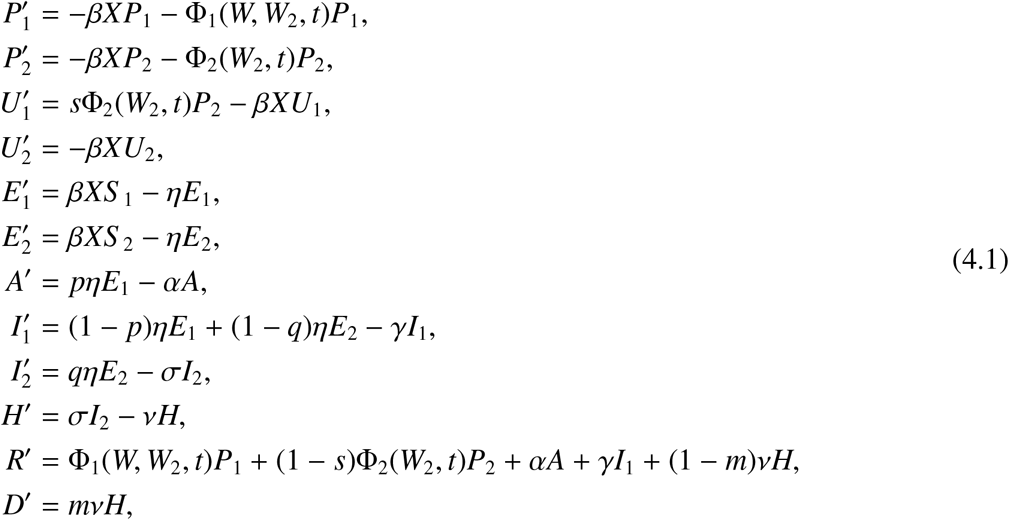

where *S* = *P* + *U*. We also need to define the quantity *X* that represents the number of individuals of class *I* needed to match the total infectivity of the actual population distribution and the functions Φ _*j*_ that define the relative rates of the vaccination processes.

#### 4.1.1. Effective infectivity *X*

Specification of *X* is based on a number of additional assumptions:

1. Fractions *c*_*i*_ of class I and *c*_*a*_ of class A are identified by a positive test. These confirmed cases have decreased infectivity because they put themselves into isolation.
2. The infectivity of each unconfirmed symptomatic infective is 1 (without loss of generality because there is the additional rate constant *β* in the transmission rate formula).
3. Asymptomatics and confirmed infectives have infectivities of *f*_*a*_ and *f*_*c*_ (both less than 1) relative to that of unconfirmed symptomatic infectives. Hospitalized infectives do not transmit the disease to an extent that justifies inclusion in the model.
4. There is a ‘contact factor’ *δ* that represents the level of risk from the average person’s sum total of encounters, relative to normal. This parameter can be used to represent both physical distancing, which decreases the rate of encounters, and wearing of masks, which decreases the risk of each encounter. It is applied to unconfirmed infectives, both symptomatic and asymptomatic, but not to confirmed infectives.

With these assumptions, the effective number of infectives is

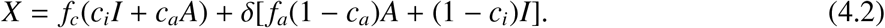

#### 4.1.2. A two-risk-class supply-and-distribution-limited vaccination model

With the population divided into low risk and high risk classes (subscripts 1 and 2, respectively), our best fit model for the population of vaccine acceptors from before (*W* from (3.5) or (3.7)) should still apply to the sum *W* = *W*_1_ + *W*_2_; we need only prescribe a model for *W*_2_ and then calculate Φ_1_ from 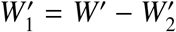. The simplest assumption is that the same model holds for *W*_2_ as for *W*, with the same parameter *K*:

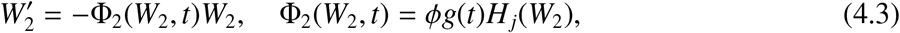

where *j* is 2 or 3.

While we do not need to keep track of *W*_1_ as a differential equation state variable, we do need to determine Φ_1_ from

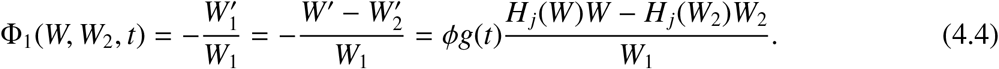

Using the formulas for *H*_*j*_, we obtain

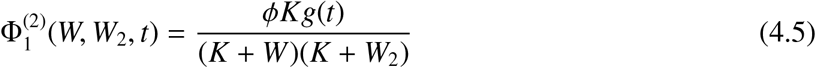

for type 2 and

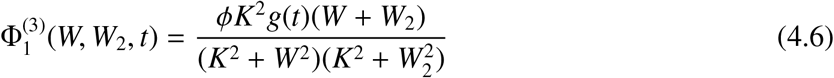

for type 3. To complete the vaccination models, we assume a fraction *k* of the population falls into the high-risk category and that the vaccine non-acceptance fractions are *r* for the general population and *r*_2_ < *r* for the high-risk subpopulation. Initial conditions are then

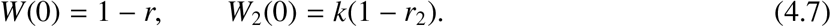

Figure 9 compares the two-class vaccination models with Holling type 2 and type 3 dynamics. There is no suitable data that separates vaccination for high-risk and low-risk classes, particularly since risk levels do not exactly correspond to CDC priority classes. However, there are qualitative differences that can help us choose between the two models; in particular, the type 2 model shows a higher efficiency in directing vaccination to high-risk individuals than the type 3. Some inefficiency clearly occurred anecdotally, as the author’s limited acquaintances include several low-risk individuals who were vaccinated early because some health-care facilities chose to vaccinate people of any risk class rather than waste defrosted doses. The type 2 model shows high-risk vaccination to have been completed within 4 months, a claim that is likely overoptimistic; thus, the type 3 model seems slightly better.

**Figure 9.**
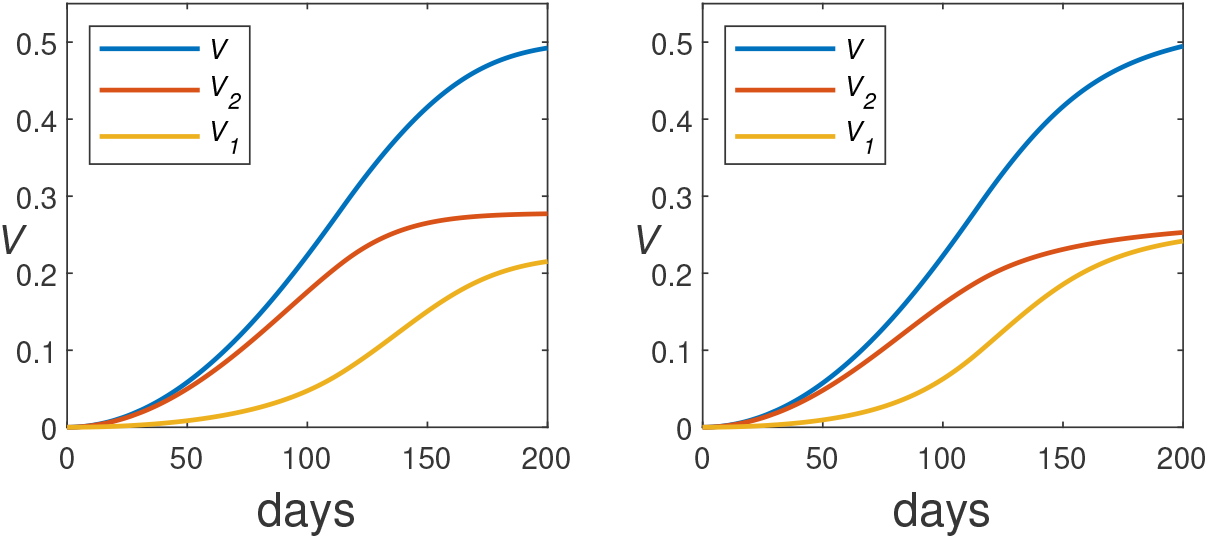
The two-class vaccination model using (3.4) for *W* and *W*_2_ (left) and (3.6) (right) and initial conditions (4.7), with *k* = 0.333, parameters *ϕ, r, K*, and *τ* from the best fits for *V* (3.10–3.11), and *r*_2_ = *r/*3. The plots show the vaccinated fractions *V*_*j*_ = *W*_*j*_(0) − *W*_*j*_.

### 4.2. Numerical Experiments

To test the impact of vaccination for a critical case, we consider a hypothetical scenario in which vaccination begins in the United States in September 25, 2020, at a period of low chronic incidence just prior to a significant spike in cases, rather than starting 3 months later in the middle of a spike, as actually happened. Parameters are listed in Table 1. The notes column refers to the following list of comments:

1. Primary value fit from US data, as shown above. Secondary values are either optimistic guesses for countries with less vaccine non-acceptance or optimistic guesses for vaccine supply in future pandemics.
2. These are guesses for parameters that cannot be measured or are strongly dependent on region or jurisdiction. The full set of these parameters has been tuned so that the results for the no vaccination case (the red curve in Figure 10) roughly match the actual results for new hospitalizations [7], which should be the quantity *σI*_2_. This quantity is initially given per 100,000 people as the parameter 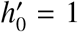 and shows a peak in the dataset of about 5 times that value after about 110 days.
3. Values for *r*_2_ are 1/3 of the values for *r* in any simulation. This assumption is suggested by US age-stratified vaccination data [6].
4. These parameter values are calculated by combining *k* with overall population estimates of 40% asymptomatic, 3% hospitalized, and 10% infection probability for vaccinated patients.
5. A large range of values of ℛ_0_ has been reported for the ancestral variant, with values in the range [2.5, 3] most common. These are clearly too low, as they cannot account for the growth of hospitalizations in March 2020 [7] nor the observation that masking and social distancing greatly reduced the incidence of seasonal flu and rhinoviruses (having ℛ_0_ < 3), without keeping COVID under control.^§^ The best estimate for the ancestral variant is ℛ_0_ = 5.8 for the United States and slightly less for Europe [16]. Because of the large spike in cases for the time period of our scenario, it seems probable that the basic reproduction number at this time had increased, either because a more infectious variant was already taking over or because of seasonal changes in contact patterns.

**Table 1.** Parameter Values for PUEAIHRD simulation

| parameter | meaning | value | ref's | notes |
| --- | --- | --- | --- | --- |
| $\mathcal{R}_0$ | basic reproduction number | 7 | | 5 |
| $h'_0$ | initial hospitalization rate per 100K | 1 | [7] | 2 |
| $r_0$ | initial $R$ | 0.3 | | 2 |
| $c_a$ | confirmed fraction for $A$ | 0.2 | | 2 |
| $c_i$ | confirmed fraction for $I$ | 0.7 | | 2 |
| $f_a$ | relative infectivity of $A$ | 0.75 | [5] | |
| $f_c$ | relative infectivity in isolation | 0.1 | | 2 |
| $k$ | high-risk fraction | 0.333 | | 2 |
| $K$ | vaccination semi-saturation const. | 0.137 | | 1 |
| $m$ | deaths per $H$ | 0.2 | [9] | |
| $p$ | low-risk asymptomatic fraction | 0.6 | [5] | 4 |
| $q$ | high-risk prehospitization fraction | 0.06 | [5, 9] | 4 |
| $r$ | overall vaccine non-acceptance probability | 0.45, 0.15 | | 1 |
| $r_2$ | high-risk vaccine non-acceptance probability | 0.15, 0.05 | | 3 |
| $s$ | breakthrough probability for high risk | 0.3 | | 4 |
| $\alpha$ | clearance rate for $A$ (1/days) | 0.125 | [3] | |
| $\gamma$ | clearance rate for $I_1$ (1/days) | 0.1 | [14, 23] | |
| $\delta$ | contact factor | 0.5 | | 2 |
| $\eta$ | clearance rate for $E$ (1/days) | 0.2 | [17] | |
| $\nu$ | clearance rate for $H$ (1/days) | 0.1 | [21] | |
| $\sigma$ | clearance rate for $I_2$ (1/days) | 0.333 | [11] | |
| $\tau$ | time to maximum vax. supply | 115, 60 | | 1 |
| $\phi$ | vaccination rate constant | 0.0057 | | 1 |

**Figure 10.**
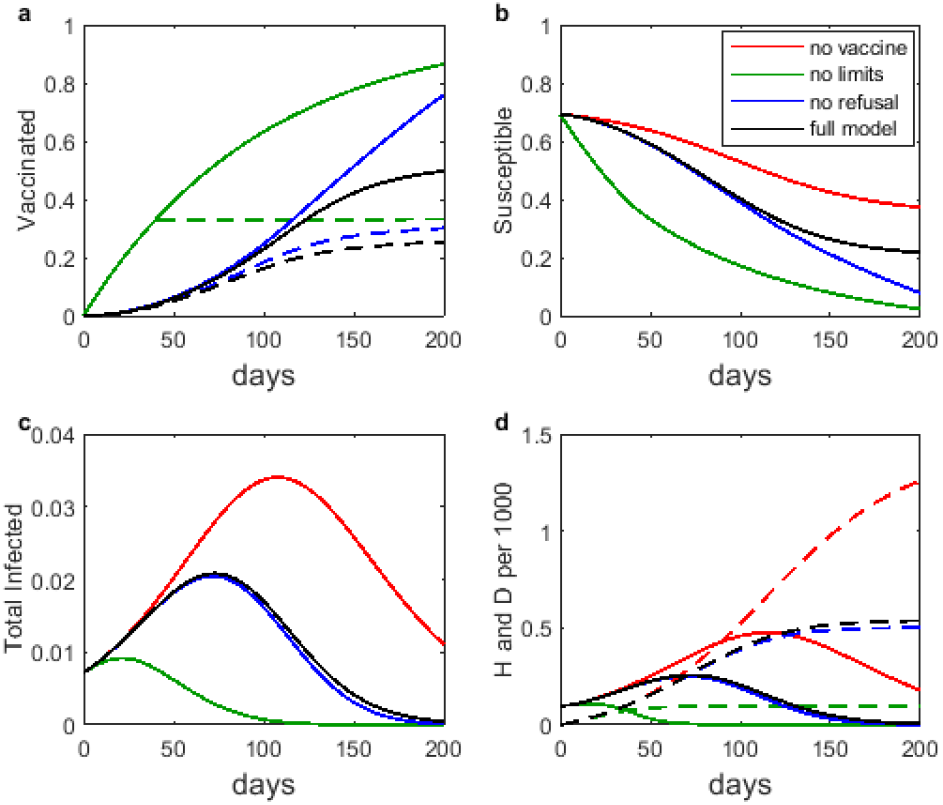
The PUEAIHRD model with various vaccination submodels: no vaccine, spontaneous transition, Holling type 3 with no vaccine non-acceptance, and the full Holling type 3 model; (a): vaccinated fraction (solid) and high-risk vaccinated fraction; (b) total susceptible fraction; (c) total infected fraction; (d) hospitalized (solid) and deceased per thousand.

#### 4.2.1. Impact of vaccination model selection

Figure 10 shows the results of an experiment to test the effects on model behavior of four different vaccination model choices:

1. No vaccination;
2. The naive spontaneous transition model with the additional naive assumption that all high-risk individuals are vaccinated before any low-risk individuals, using *ϕ* = 0.01, corresponding to a 100-day mean time for vaccination;
3. The two-class Holling type 3 vaccination model (3.6, 4.3) with best-fit parameters for *ϕ, K*, and *τ*, but neglecting vaccine non-acceptance;
4. The two-class Holling type 3 vaccination model with best-fit values for all parameters.

The no-vaccine scenario shows a significant wave of cases, as happened in the actual event, with correspondingly high hospitalizations and deaths. The naive vaccination model shows vaccination having a dramatic impact on this wave; however, the vaccination plot in panel **a** shows how unrealistic this is. With a better vaccination model, we see that vaccination would have had a significant positive impact, but much less than the impact seen with the naive model.

The impact of vaccine non-acceptance is larger here than in the PUEIR model example because the prioritization of high-risk individuals makes vaccine non-acceptance significant earlier in the scenario. The difference between the full acceptance and the realistic acceptance results for vaccine doses administered to high-risk individuals becomes noticeable on the panel **a** graph at about 70 days. This is not enough to significantly affect the total number infected in panel **c**, but it does make a visual difference in the hospitalization and death counts of panel **d**. Future spikes for scenarios starting after the displayed one will play out quite differently in these two cases because the total susceptible population drops to a far greater extent with full vaccine acceptance (panel **b**).

#### 4.2.2 Impact of vaccination model parameters

We now use the PUEAIHRD epidemic model with the two-class supply-limited Holling type 3 vaccination model to explore the impact of several key parameters that can be influenced by public policy decisions. Figure 11 shows the results. There are six combinations of assumptions:

1. The default scenario, in which there is no vaccination;
2. The base scenario, in which vaccination parameters are from the best-fit values and the contact factor is *δ* = 0.5;
3. The enhanced acceptance scenario, in which vaccine non-acceptance is reduced by a factor of 3, equivalent to replacing the United States with one of the countries having highest vaccine acceptance in 2021;
4. The enhanced manufacture scenario, in which advance preparation reduces the time required for vaccination to reach full capacity from 115 days to 60 days;
5. The combined enhancement scenario, combining reduced vaccine non-acceptance with faster achievement of capacity;
6. The enhanced mitigation scenario, in which vaccine parameters are not enhanced, but the contact factor is reduced from 0.5 to 0.4.

**Figure 11.**
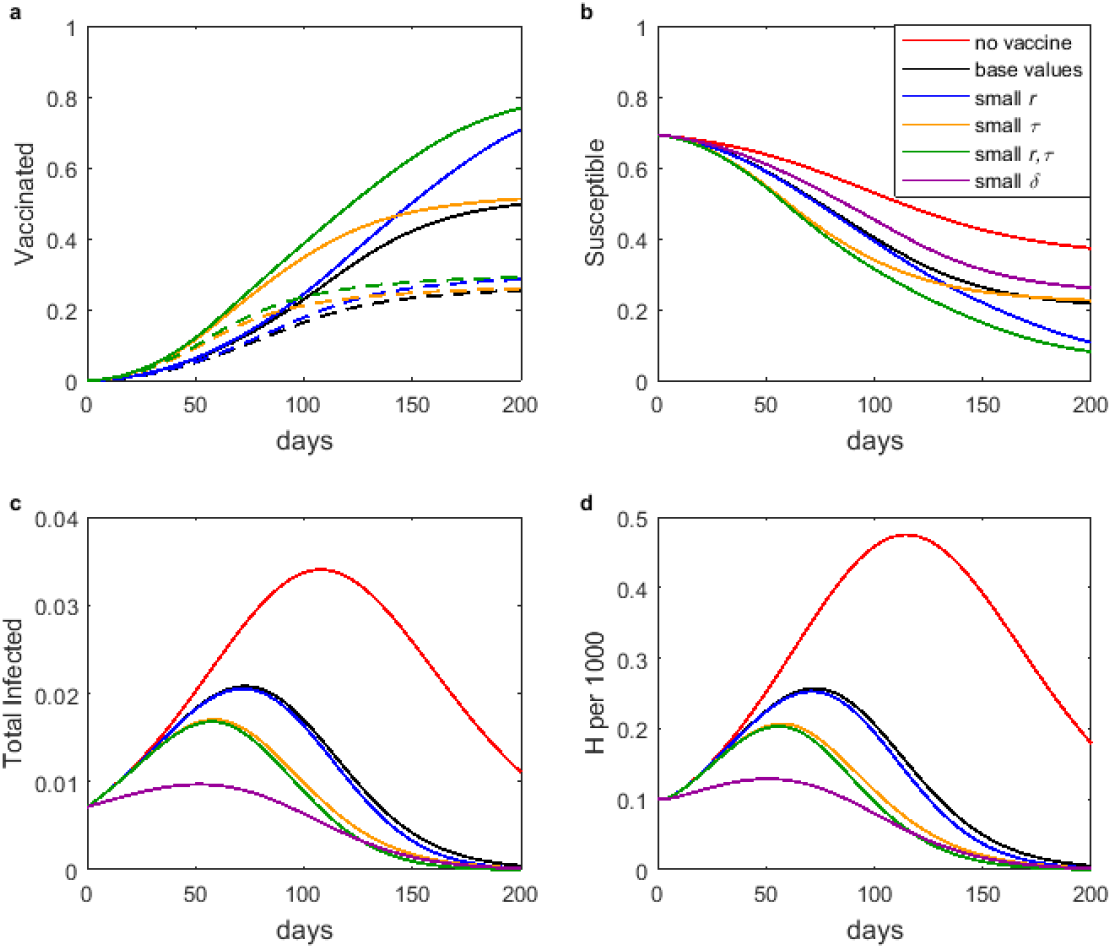
The PUEAIHRD model with various assumptions about vaccination and mitigation: no vaccine, base parameter set, less non-acceptance, faster rollout, less non-acceptance and faster rollout, base parameter set with lower contact factor; (a): vaccinated fraction (solid) and high-risk vaccinated fraction; (b) total susceptible fraction; (c) total infected fraction; (d) hospitalized per thousand.

Panel **a** shows the vaccination outcomes for the different scenarios. Enhanced acceptance raises the overall vaccination numbers at the end of the scenario, while enhanced manufacturing leads to an earlier approach to those maxima. The curve for scenario 6 is omitted, as it is the same as that for scenario 2.

Panel **b** shows the time history of the total susceptible population. Of primary interest are the values at the end of the scenario, as these values are predictors of the severity of the next outbreak of the same disease. Here we see the value of enhanced vaccine acceptance.

Panels **c** and **d** show the time histories of the total infected and hospitalized. Enhanced vaccine acceptance makes only a small difference in these outcomes, while enhanced ramping speed makes a much larger difference. The effect of a modest improvement in the standard mitigation practices of social distancing and masking is larger than the impact of enhancements to vaccination, however.

The graphs of total cases and total hospitalized are almost identical aside from the scale, except for one noticeable difference. In general, vaccination has a greater effect on hospitalization than it has on infection in our COVID-19-based scenario; this is clearly due to a combination of prioritization of high-risk individuals for vaccination combined with the large probability that vaccination of these individuals reduces the severity of the disease without reducing the incidence.

## 5. Discussion and Conclusions

COVID-19 has provided an ideal opportunity to develop modeling tools that can be adapted to new diseases. One area where previous models fall short is the incorporation of vaccination subject to vaccine non-acceptance and limitations on supply and distribution. The models developed in this paper will likely be suitable for novel disease pandemics of the future. They are more complicated than the standard single-phase transition model, so it is important to be clear about the benefits.

### 5.1. Vaccine Non-Acceptance

As of April 2022, the fractions of national populations that have received a full initial vaccine protocol (not counting boosters) was as high as 96% in the United Arab Emirates, but had a worldwide average of only 59% [28]. The actual percentages of people who have been fully protected at any given time are surely smaller, as the initial protocol without boosters was no longer adequate in April 2022. In some cases, the main difficulty is global inequity, but in other cases, such as the United States (just 66% of people having had the full initial protocol), the problem is a combination of slow approval of vaccination for children and adolescents, resistance to vaccination, and hesitancy. These reasons for non-acceptance will likely be present for novel diseases of the future, so it is important to account for it in models. This requires models that partition the standard susceptible class into prevaccinated and unprotected classes. Of course the extra state variable adds additional complexity to a model, which should be avoided when the extra detail is not necessary.

For an endemic disease, the benefits of vaccination can be significantly reduced by widespread vaccine non-acceptance, as illustrated in Figures 2 and 3, particularly for diseases where immunity is short-lived and vaccination must be renewed regularly. Here, vaccine resistance will be augmented by apathy among people who lose interest in renewing their immunity. Vaccine non-acceptance can significantly reduce the public health benefits of vaccination for diseases that are highly transmissible and have immunity that lasts only a few years or less.

Vaccine non-acceptance is largely irrelevant in many vaccine rollout scenarios, as there are not enough doses at first to fully accommodate those who want to be vaccinated. As more doses become available and more people are vaccinated, vaccine non-acceptance begins to have an impact. As shown in Figures 10 and 11, vaccine non-acceptance by high-risk individuals makes a noticeable difference in hospitalizations and deaths, although neither vaccine non-acceptance in general nor that specifically for high-risk individuals makes much difference to the overall course of the pandemic.

### 5.2. Supply and Distribution

Supply and distribution issues are critical in epidemic scenarios, when there is a large queue of people wanting vaccination and limited numbers of doses and appointment slots. The significant difference between the outcomes of the naive vaccination model and more realistic models, as seen in Figure 10, clearly shows the necessity of using a realistic vaccination model.

In terms of good public health outcomes, it is more important in the epidemic stage of a disease to have a good infrastructure for manufacture and administration than it is to increase vaccine acceptance, as seen in the better performance of scenarios with enhanced rollout speed compared to scenarios with enhanced acceptance in Figure 11. By the time a disease has become endemic, however, most people only need vaccination when they are due for a booster, so we can expect doses and appointment slots to be adequate. Endemic disease scenarios do not require the more sophisticated accounting for supply and distribution that is needed for epidemic scenarios.

### 5.3. Maintaining Mitigation

The comparison of scenarios in Figure 11 shows that even with the most optimistic vaccination parameters, the effect of a small decrease in contact factor is greater than the effect of vaccination by itself. While quickly eliminating unpopular mitigation practices makes for good politics, it is much better for public health outcomes to delay elimination of these practices until vaccination has become widespread enough to make a difference. This observation is self-evident, but modeling such as has been done here serves to offer confirmation and quantitative estimates.

## Data Availability

All data produced in the present work are contained in the manuscript

## Appendix – Proof of Theorem 1

For simplicity, we omit terms of *O*(*ϵ*) from the proof.

Claim 1^*^

The disease-free equilibrium was calculated in Section 2, but it remains to determine its stability. Eliminating *R* from the first three equations of (2.7) yields

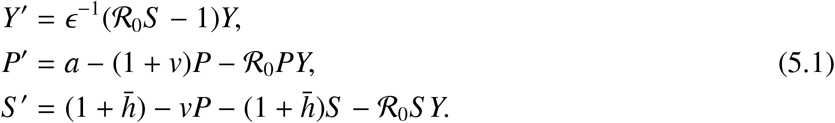

The Jacobian for this general system is

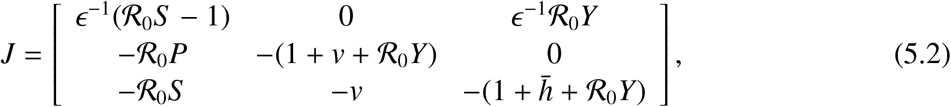

and the Jacobian at the disease-free equilibrium (0, *P*_0_, *S* _0_) is

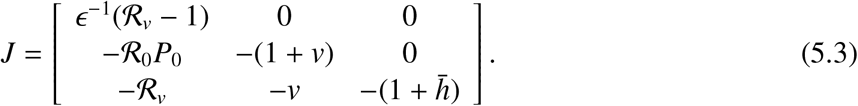

This system decouples, with two negative eigenvalues and the third given by *ϵ*^−1^(R_*v*_ − 1); hence, the disease-free equilibrium is asymptotically stable if and only if R_*v*_ < 1.

Claim 3*

Any endemic disease equilibrium must have

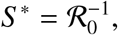

so the corresponding Jacobian is

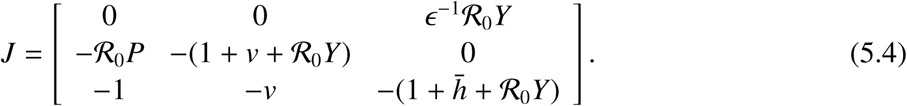

The coefficients of the characteristic polynomial *λ*^3^ + *c*_1_*λ*^2^ + *c*_2_*λ* + *c*_3_ are

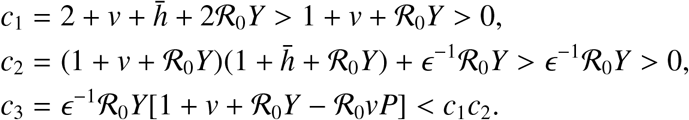

The requirement 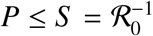 means that 1 + *v* + ℛ_0_*Y* ℛ_0_*vP* ≥ 1 + ℛ_0_*Y >* 0. Thus, the Routh-Hurwitz conditions [18, 19] are satisfied, so any endemic equilibrium point must be asymptotically stable.

Claim 2

The endemic disease equilibrium values *Y*^*^ and 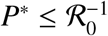 are defined by the system

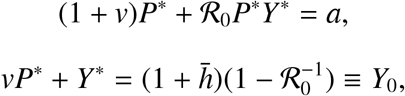

Substituting from the second equation into the first gives the quadratic equation

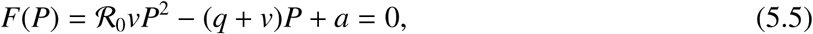

where

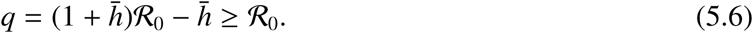

This function satisfies the properties

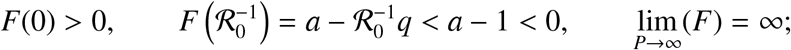

hence, *F* = 0 has two real roots that lie on either side of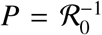. This means that there is always a unique solution with 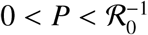 that is given by

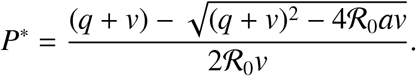

Associated with this solution is the unique value *Y*^*^ given by (2.10).

We have now shown that the equilibrium equation (5.5) always has a unique solution with *P* in the acceptable range, but this solution is meaningful only if *Y*^*^ *>* 0. To discover the existence condition for the endemic disease equilibrium, we assume by way of contradiction that *Y*^*^ ≤ 0. Then the second term in the numerator of the formula for *Y*^*^ must be negative and larger in magnitude than the first term, which is always positive. Hence,

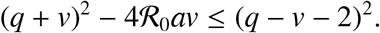

Simplification of this inequality eventually yields

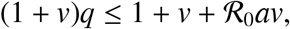

which we can manipulate to get ℛ_*v*_ ≤ 1. Hence, we get an unacceptable solution with *Y*^*^ ≤ 0 whenever ℛ_*v*_ ≤ 1; conversely, we get an acceptable solution with *Y*^*^ *>* 0 whenever ℛ_*v*_ *>* 1.

© 2022 the Author(s), licensee AIMS Press. This is an open access article distributed under the terms of the Creative Commons Attribution License (http://creativecommons.org/licenses/by/4.0)

## Footnotes

† The total susceptible population *S* figures prominently in the equilibrium and stability analysis.

‡ The state variables are already scaled by the total population.

§ While the SIR model is a good base model for endemic settings, the fact that the time scale for incubation is roughly comparable to that for infectious duration means that the SEIR model is a better base model for epidemic settings.

¶ This figure represents the fraction of the population that has become immune through infection prior to the start of the scenario.

* We are assuming that exposure risk is not considered in vaccination policies. Including this feature would require many more classes and heterogeneous mixing [10]. In the United States, most jurisdictions prioritized health-care workers, which constitute only a small fraction of the total population, but not workers in other areas deemed essential, such as food processing.

† The classification of individuals as high or low risk is imprecise at best, so there is no loss of generality in assuming that anyone who becomes asymptomatic must have been low risk, while anyone who has a breakthrough infection after vaccination or becomes hospitalized must have been high risk.

‡ With these assumptions we can control the extent to which vaccination only reduces severity rather than inducing full immunity. As in the discussion of *E*_1_ and *E*_2_, the lack of precision in categorizing risk levels allows us to assume that all breakthrough cases were among high-risk individuals. The incidence of hospitalization for fully immunized individuals was low enough to justify neglecting that possibility in a relatively simple model.

§ Global market reports for over-the-counter cold remedies show a significant decline in the winter of 2020-2021 in spite of the use of these medications by COVID patients [8]. The only explanation is that incidence of seasonal rhinovirus was much less than in the prior year.

* We consider claim 3 before claim 2, as its proof is straightforward. As is often the case with simple models, the proof of stability of endemic disease equilibria for the PUIRU model does not require formulas for the equilibria or conditions for existence and uniqueness.

## References

1. J. Arino, S. Portet, A simple model for COVID-19, Infectious Disease Modelling, 5 (2020), 309–315, DOI: 10.1016/j.idm.2020.04.002.

2. F. Brauer, C. Castillo-Chavez, Z. Feng, Mathematical Models in Epidemiology. Springer-Verlag, New York, 2019.

3. A.W. Byrne, D. McEvoy, A.B. Collins, K. Hunt, M. Casey, A. Barber, F. Butler, J. Griffin, E.A. Lane, C. McAloon, K. O’Brien, P. Wall, K.A. Walsh, S.J. More, Inferred duration of infectious period of SARS-CoV-2: rapid scoping review and analysis of available evidence for asymptomatic and symptomatic COVID-19 cases, BMJ Open, 10 (2020), https://doi.org/10.1136/bmjopen-2020-039856.

4. L.-M. Cai, Z. Li, X. Song, Global analysis of an epidemic model with vaccination, J. Appl. Math. Comp., 57 (2018), 605–628, https://doi.org/10.1007/s12190-017-1124-1.

5. The Centers for Disease Control and Prevention, COVID-19 Pandemic Planning Scenarios, 2020. Available from https://www.cdc.gov/coronavirus/2019-ncov/hcp/planning-scenarios.html#five-scenarios.

6. The Centers for Disease Control and Prevention, COVID-19 Vaccinations in the United States, 2022. Available from https://covid.cdc.gov/covid-data-tracker/#vaccinations_vacc-total-admin-rate-total.

7. The Centers for Disease Control and Prevention, New Admissions of Patients with Confirmed COVID-19, United States, 2022. Available from https://covid.cdc.gov/covid-data-tracker/#new-hospital-admissions.

8. Cough and Cold Preparations Global Market Report 2021: COVID-19 Implications and Growth to 2030. The Business Research Company, 2021.

9. The COVID Tracking Project. Our Data, 2020. Available from https://covidtracking.com/data.

10. E.H. Elbasha, A.B. Gumel, Vaccination and herd immunity thresholds in heterogeneous populations, J Math Biol (2021), doi: 10.1007/s00285-021-01686-z.

11. C. Faes, S. Abrams, D. Van Beckhoven, G. Meyfroidt, E. Vlieghe, N. Hens, Time between symptom onset, hospitalisation and recovery or death: statistical analysis of Belgian COVID-19 patients. Int J Environ Res and Public Health, (2020), doi: 10.3390/ijerph17207560.

12. S. Gazit, R. Shlezinger, G. Perez, R. Lotan, A. Peretz, A. Ben-Tov, E. Herzel, H. Alapi, D. Cohen, K. Muhsen, G. Chodick, T. Palaton, SARS-CoV-2 naturally acquired immunity vs. vaccine-induced immunity, reinfections versus breakthrough infections: a retrospective cohort study Clin Infect Dis (2022), doi: 10.1093/cid/ciac262.

13. S. Greenhalgh, C. Rozins, A generalized differential equation compartmental model of infectious disease transmission, Infectious Disease Modelling, 6 (2021), 1073–1091, doi: 10.1016/j.idm.2021.08.007.

14. X. He, E.H.Y. Lau, P. Wu, X. Deng, J. Wang, X. Hao, Y.C. Lau, J.Y. Wong, Y. Guan, X. Tan, X. Mo, Y. Chen, B. Liao, W. Chen, F. Hu, Q. Zhang, M. Zhong, Y. Wu, L. Zhao, F. Zhang, B.J. Cowling, F. Li, G.M. Leung, Temporal dynamics in viral shedding and transmissibility of COVID-19, Nat Med, 26 (2020), 672–675, doi: 10.1038/s41591-020-0869-5.

15. 15. H.W. Hethcote, The mathematics of infectious diseases. SIAM Rev, 42 (2000), 599–653.

16. R. Ke, E. Romero-Severson, S. Sanche, N. Hengartner. Estimating the reproductive number ℛ_0_ of SARS-CoV-2 in the United States and eight European countries and implications for vaccination. J. Theo. Biol., 517 (2021), doi: 10.1016/j.jtbi.2021.110621.

17. S.A. Lauer, K.H. Grantz, Q. Bi, F.K. Jones, Q. Zheng, H.R. Meredith, A.S. Azman, N.G. Reich, J. Lessler, The incubation period of coronavirus disease 2019 (COVID-19) from publicly reported confirmed cases: estimation and application. Annals of Internal Medicine, (2020), doi: 10.7326/M20-0504.

18. G. Ledder, Mathematics for the Life Sciences: Calculus, Modeling, Probability, and Dynamical Systems, Springer-Verlag, New York, 2013.

19. G. Ledder, Mathematical Modeling for Epidemiology and Ecology, 2nd edition, Springer-Verlag, New York, in press.

20. G. Ledder, M. Homp, Using a COVID-19 model in various classroom settings to assess effects of interventions, PRIMUS, 32 (2021), 278–297, DOI: 10.1080/10511970.2020.1861143.

21. J.A. Lewnard, V.X. Liu, M.L. Jackson, M.A. Schmidt, B.L. Jewell, J.P. Flores, C. Jentz, G.R. Northrup, A. Mahmud, A.L. Reingold, M. Petersen, N.P. Jewell, S. Young, J. Bellows, Incidence, clinical outcomes, and transmission dynamics of severe coronavirus disease 2019 in California and Washington: prospective cohort study, BMJ, 369 (2020), doi: 10.1136/bmj.m1923.

22. K. Liu, Y. Lou, Optimizing COVID-19 vaccination programs during vaccine shortages, Infectious Disease Modelling 7 (2022), 286–298, doi: 10.1016/j.idm.2022.02.002.

23. Y. Liu, L.-M. Yan, L. Wan, T.-X. Xiang, A. Le, J.-M. Liu, M. Peiris, L.L.M. Poon, W. Zhang, Viral dynamics in mild and severe cases of COVID-19, Lancet Infect Dis 20 (2020), 656–657, doi: 10.1016/S1473-3099(20)30232-2.

24. N.E. MacDonald, SAGE Working Group on Vaccine Hesitancy, Vaccine hesitancy: definition, scope and determinants, Vaccine 33 (2015), 4161–4164, doi: 10.1016/j.vaccine.2015.04.036.

25. M. Martcheva, An Introduction to Mathematical Epidemiology. Springer-Verlag, New York, 2015.

26. A.R. McLean, S.M. Blower, Imperfect vaccines and herd immunity to HIV, Proc. R. Soc. Lond. B, 253 (1993), 9–13.

27. S.M. Moghadas, T.N. Viches, K. Zhang, C.R. Wells, A. Shoukat, B.H. Singer, L.A. Meyers, K.M. Neuzil, J.M. Langley, M.C. Fitzpatrick, A.P. Galvani, The impact of vaccination on coronavirus disease 2019 (COVID-19) outbreaks in the United States, Clin Infect Dis, 73 (2020), 2257–2264, doi: 10.1093/cid/ciab079.

28. Our World in Data, Coronavirus Vaccinations, 2022. Available from https://ourworldindata.org/covid-vaccinations.

29. Our World in Data, State-by-state Data on COVID-19 Vaccinations in the United States, 2022. Available from https://ourworldindata.org/us-states-vaccinations.

30. P. van den Driessche, J. Watmough, Reproduction numbers and sub-threshold endemic equilibria for compartmental models of disease transmission, Math. Biosci., 180 (2002), 29–48, doi: 10.1016/S0025-5564(02)00108-6.

